# Improving Nutritional Status Among Moderately Underweight Infants in Mumbai; Results of a Randomised Controlled Trial to Evaluate *Swasth Kadam*, an mhealth Intervention

**DOI:** 10.64898/2025.12.10.25342028

**Authors:** Neha Madhiwalla, Hemlata Jiwnani, Hardik Bhatt, Bharati Jadhav, Aparna Hegde

## Abstract

This paper reports the results of a randomised controlled trial **(CTRI/2023/01/048939)** conducted to evaluate the outcomes of a telephonic counselling service for mothers of moderately underweight infants aged 6-24 months as defined by the WHO standards and residing in low-income settlements of the Mumbai metropolitan region (N=713). The primary outcome was accelerated weight gain and reduction in the decline to more severe levels of malnutrition. These were measured in terms of the change in the percentile category between baseline and endline using the standard WHO tables. Results indicate that the proportion of infants who showed no change or deterioration in the percentile category of weight was higher in the control group. Moreover, analysis of disaggregated data indicate that the gains of the intervention were more significant for more vulnerable infants such as those in the lowest percentile of weight at baseline (Chi square = 6.786; p = 0.034), born with low birth weight (Chi square = 8.402; p = 0.015), girls (Chi square = 6.590; p = 0.037), and infants whose fathers were in unskilled occupations (Chi square = 6.033; p = 0.049). Despite the limitations of sample size and being a single site study, these results point to the potential of an mhealth intervention to address the widespread problem of malnutrition among infants aged 0-2 years, particularly among vulnerable groups, which are unable to access actionable information and advice from other sources.

## INTRODUCTION

Undernutrition is one of the major problems affecting child health in the global South (1,2). both in rural and urban areas (3). In India, the latest estimates from 2019-21 indicate that 27 percent of urban children and 34 percent of rural children under the age of five years are underweight (4). With growth in urbanization in India and consequent increase in urban poverty, childhood malnutrition is widely prevalent in urban slums (5,6,7). Two thirds of these children suffer from anaemia. Supplementary nutrition and growth monitoring through community-based centres remain the key interventions to address this problem. Complementally, counselling to influence feeding, health and hygiene practices can have a significant impact on recovery (8,9,10). Several models of maternal counselling have been developed and implemented in different settings (11,12). A randomised controlled trial conducted to evaluate home based growth monitoring and counselling reported an increase in height-for-age z score and also for weight-for-age z scores in the age group between 3 to 21 months (13). There are few studies that elicit improvement in anthropologic measurements. A small study in India (N =57 mothers) showed marginal improvement in the protein intake and weight in the intervention group (14).

Given the magnitude of the problem, it is logistically challenging to deliver complex health advice and support within public health systems that depend on lay health workers to scale up programmes (15). These workers do not have required skills to engage with the caregiver in problem-solving (16). Additionally, mothers’ perception or lack of support hinders uptake of community-based interventions (17).

Remote solutions offer the promise of a scalable solution to overcome the gap in expertise which hamper such interventions. mHealth interventions which have been implemented in this domain include the use of digital tools devices for measurement (18,19), job aids of health workers to facilitate data collection and surveillance (20,21), health messaging via sms, audio and video either directly delivered to the end users or mediated by a healthcare worker. Both one way and two-way communication modalities have been tested in diverse settings (22). However, there is very little evidence of large public health mhealth programmes that show effective outcomes (23).

Teleconsulting or telehealth, encompassing patient education, health-care related information by trained professionals (24) has seen a major rise during Covid19 pandemic and has proven to be an important bridge in absence of sufficient health services. Tele counselling services as part of programmes for nutrition education have proven to be effective resulting in improved breastfeeding and increased weight gain in subsequent visits by mothers who received mobile based counselling (24, 25) albeit with limitations in sample size and generalisability.

Nutritional counselling has been found to improve dietary intake in children (26, 27). There was a paucity of literature available to start the intervention earlier in 0–3-year age group and those who have mild or moderate malnutrition as focus of intervention for WHO and UNICEF has always been therapeutic intervention for children who have Moderate Acute Malnutrition (MAM) or Severe Acute Malnutrition (SAM). This paper presents the results of a community based randomized controlled trial for improving adherence to feeding, health and hygiene best practices among caregivers of moderately underweight children aged 6-36 months through telephonic counselling. This study was conducted in 2023-2024 in more than 20 low-income settlements in Mumbai Metropolitan Region (MMR), India.

## AIMS and OBJECTIVES

The study was designed to test a free telephonic counselling service in which trained nutrition/health counsellors interacted with caregivers of moderately underweight infants (WAZ score < -2 SD and > – 3SD) aged 6-24 months through 12 direct calls on food, health and nutrition weekly for a period of 3 months. The objectives of the study were to compare the nutritional status of infants in the control and intervention groups at the end of six months, using WHO growth standards. Secondarily, it was also aimed to compare the adherence to best practices in feeding, health and hygiene among caregivers of infants in the intervention group vs. control group at the end of six months using a set of standard parameters. In this paper we report the results pertaining to the primary outcome, accelerated weight gain measured by overall weight gain and change in the percentile category adjusted for age and sex.

**Figure 1:**
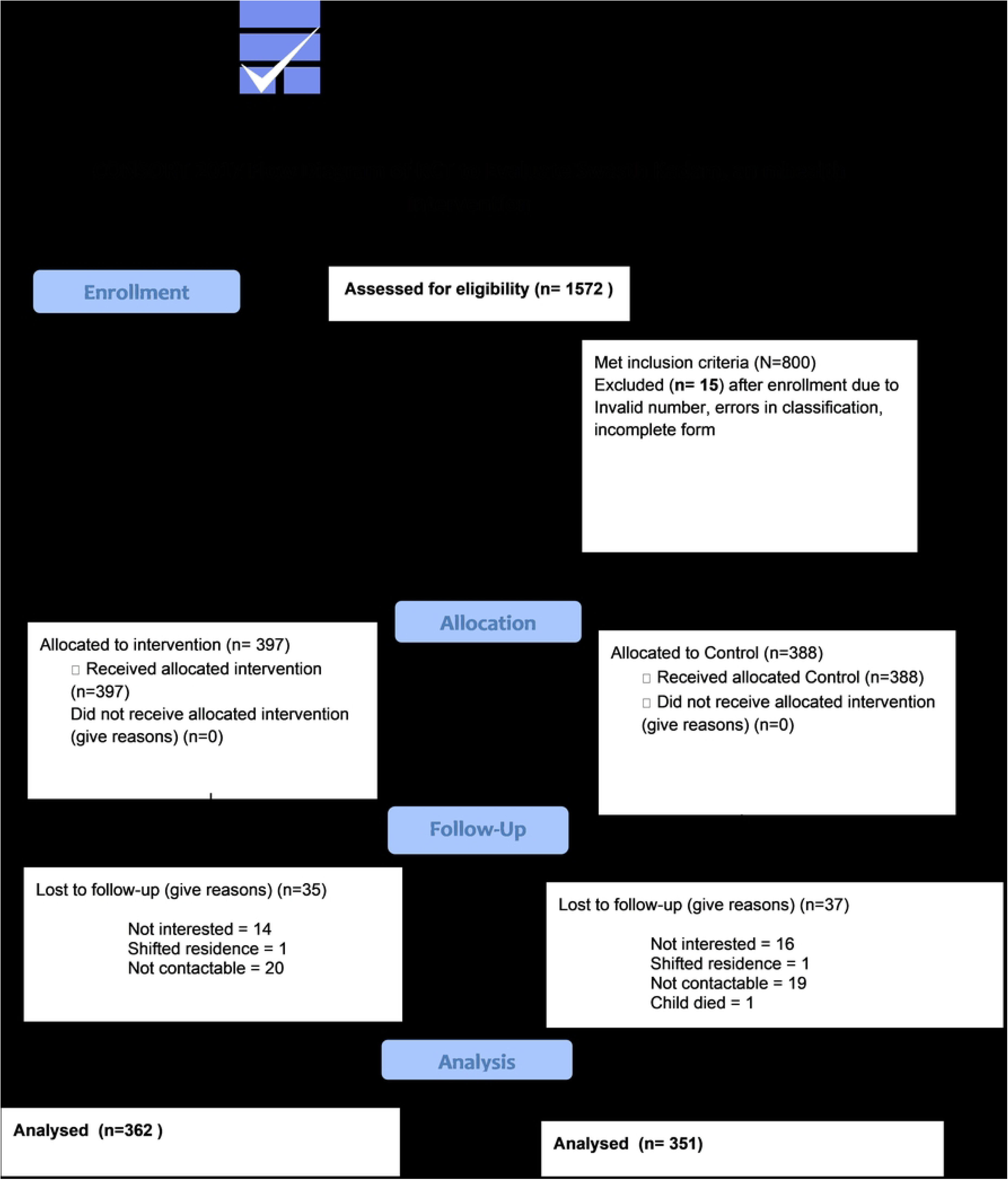
CONSORT FLOW DIAGRAM: Improving Nutritional Status Among Moderately Underweight Infants by Promoting Adherence to Best Practices in Nutrition, Health and Hygiene Among Caregivers in Mumbai.

## STUDY SETTING

The participants were recruited from more than 20 clusters of informal settlements (slums) located in the Mumbai Metropolitan Region. In general, the study population was socially disadvantaged. 43.3 percent of the mothers had not completed school. About one fourth of the women had completed 6^th^ to 9^th^ grade (25.8 percent) and 17.5 percent had even less education than these. Another one fourth had completed the 10^th^ grade (24.8 percent). The remaining had completed higher secondary education or more. The majority of the fathers were unskilled workers (78.2 percent). Only 9.7 percent of the fathers were engaged in a service sector job. Among these participants, 42 percent of the families were living in rented housing. Typically, these settlements were marked by poor environmental conditions, including poor ventilation and light inside the homes, and open drains, overflowing garbage bins and stagnant water collecting in puddles outside the houses.

## MATERIALS AND METHODS

The sample size calculation for a superiority study using two-sided test of means, assuming 20% improvement in weight gain with 80% power and 95% CI, was 784 children. To compensate for sample loss, the sample size was set at 800 children with equal allocation to both arms. The study was conducted as a single stage randomised controlled trial with intention to treat design. The study was reviewed and approved by the ARMMAN Ethics Review Board. The study was registered as an interventional trial with the Clinical Trials Registry of India (CTRI/2023/01/048939) with the title “Improving adherence to best practices in feeding, health and hygiene amongst moderately underweight children through telephonic counselling”. The baseline study was commenced on 16^th^ January 2023 and the last endline interview was conducted on 7^th^ December 2023.

As noted above, participants were recruited in high prevalence areas, which were typically low income, of the Mumbai Metropolitan Region. The WHO child growth standards *“(*https://www.who.int/tools/child-growth-standards/standards/weight-for-age*)”* were used for screening and selection for participation. After screening, all eligible infants whose primary caregiver consented to participation were recruited for the study. The inclusion criteria for the study were any child who fit the definition of moderately underweight, was aged between 6-24 months and intended to reside in this location for at least the next six months. The exclusion criteria included any child who had comorbidities requiring continuous medical monitoring. Written informed consent was sought from the mothers after offering a detailed explanation of the study design and process. All caregivers were informed that they would have a 50 percent probability of being included for counselling. Otherwise, they would be offered that service at the end of six months, if their child was still malnourished. The individual infants of all consenting participants enrolled in the preceding week were randomly allocated to intervention or control arm using computer generated random number tables. During the period of the trial, a withdrawal criterion was defined to withdraw any child, who is diagnosed with a medical condition, intervention, develops pitting oedema or has loss of appetite. Such children were referred to the virtual OPD programme of ARMMAN. The caregivers were also advised to consult a health facility. Being a minimal harm study, specific harms resulting from the intervention were less likely and pertained to actions resulting from misunderstanding advice or improper implementation, such as not maintaining hygiene during food preparation. The team would provide appropriate referral and take responsibility for treatment.

A baseline survey was conducted to record basic demographic information, the mother’s obstetric history and details of nutritional practices in the first six months of the life of the index child. At the end of the trial, an endline survey was conducted to record the child’s current weight, along with details of current dietary practices, health-seeking and childcare practices. Only those participants for whom complete data of both baseline and endline was available were included for the final analysis. For measurement of the primary outcome, a difference in difference analysis to assess the impact of the intervention on accelerated weight gain measured in terms of absolute recovery, weight being normal for age and sex (WAZ > -2SD) or decline to severe underweight (WAZ < -3SD) and relative improvement in terms of movement to a higher percentile category of weight adjusted for age and sex.

## INTERVENTION DESIGN

An amalgam of the guidelines issued by the WHO and ICDS focusing on the community care of children with moderate malnutrition were used to design the framework of the intervention. These guidelines endorse the following interventions: growth monitoring, deworming, iron supplementation, learning-by-doing sessions on feeding practices, health, hygiene and psychosocial practices, counselling on care practices, referral for routine medical services (immunisation, health check-ups) as well as medical complications. The Swasth Kadam intervention design was informed by these guidelines. Child nutrition programmes have often been criticised for deepening the burden on the mother and ignoring the role and responsibility of other family members towards caregiving (28, 29). Hence, a gender rights framework (30, 31) was used to adapt and reorient the material to enhance the participation of other family members in the care of the child and also address the mother’s own health and well-being. Thus, in addition to the above themes, we include components on maternal health, maternal stress, involvement of the family in childcare, inter-spousal communication and family planning.

Standard operating procedures pertaining to the periodicity of calls, protocol for follow-up and referral, addressing non-response and dropout were adopted to ensure that the calling was standardised. Monthly debriefing meetings were conducted to address specific health queries raised by the participants, discuss logistical and technological challenges faced by the counsellors and challenges posed by the participants’ personal and social situation. 11 debriefing meetings, each of 3 to 4 hours duration were held in the period of the study.

All enrolled participants were assigned to the counsellors in round-robin format. Typically, the same counsellor delivered all the calls to an individual participant. A standard protocol of 13 calls was set. An introductory call was made to take consent and brief about the programme. Future calls were scheduled based on mutually decided day and time as per the convenience of the participant. The day and time of the call was decided mutually. A customised web-application enabled the counsellor to schedule calls and also record feedback.

Key domains for counselling had been outlined based on the intensive formative research and programme experience. These included growth and development, breastfeeding, complementary feeding, prevention and treatment of infections, hygiene, maternal well-being, and family planning. A total of 32 sub-themes were identified within these domains. Key messages and information relevant to each sub-theme were been converted into a script. These scripts were available to the counsellor within the web-application in an easily searchable and retrievable format. The sub-themes were organised in a sequence based on a logical flow of information.

**Figure 2:**
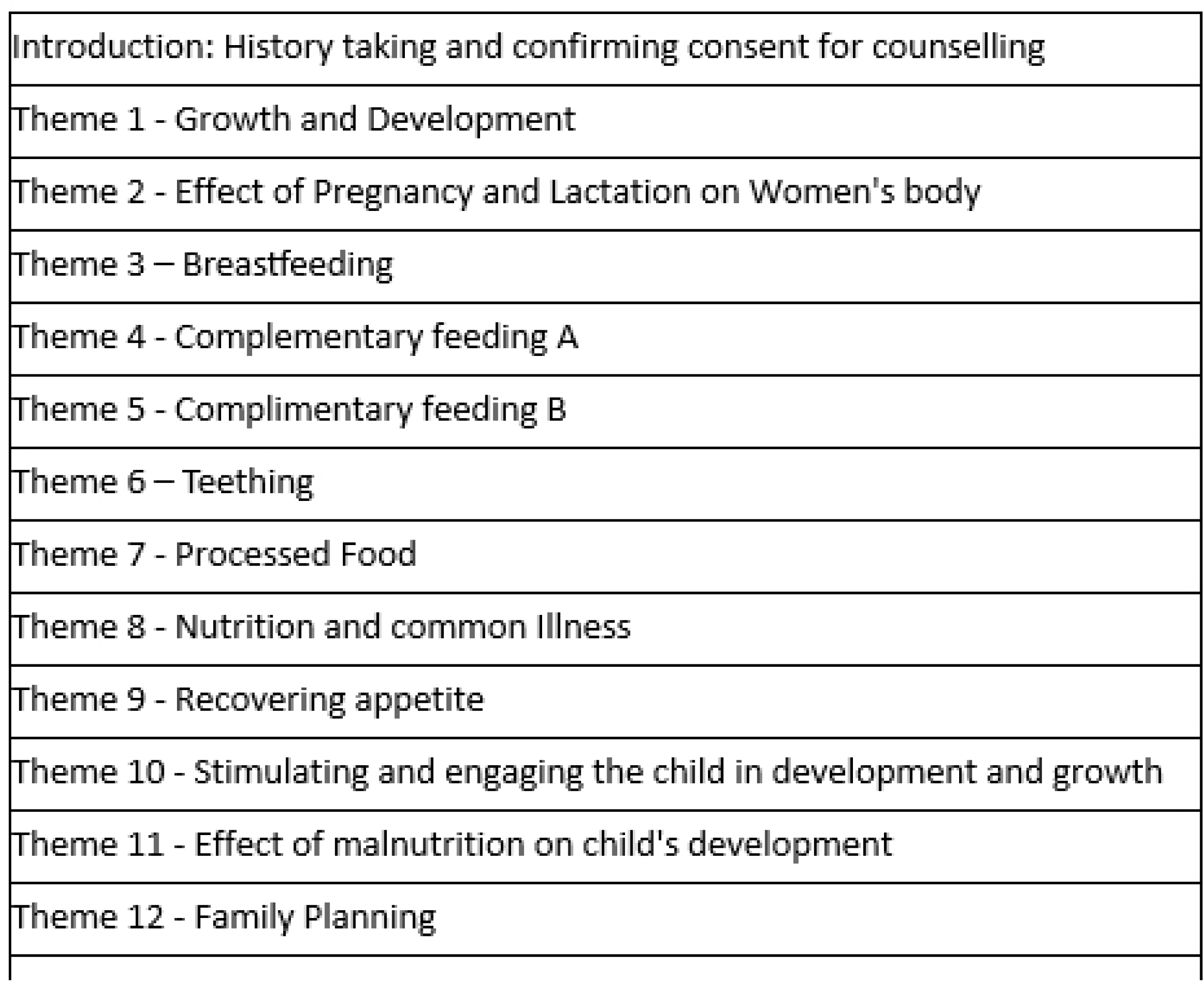
Key domains of nutrition and health information covered in the counselling calls.

Typically, the counsellor was expected to deliver content on 2-4 sub-themes in each call in the given sequence. However, counsellors could alter the sequence based on the immediate information needs of the participant or in response to an acute event (e.g. illness, loss of appetite, concern about unwanted pregnancy, etc.). In addition to the scripted content, there were extempore conversations which typically included updates on events, problems experienced in implementing advice, other health and social problems. Based on the individual needs of the caregiver, each sub-theme could be repeated multiple times.

In every call, the counsellor elicited information about the child’s weight, appetite and general health condition. A service was considered completed when all the themes/sub themes had been discussed. While a call was scheduled with a gap of seven days, it was usual for the service to take up to 4 months to be completed due to unavailability of caregiver or problems pertaining to the phone and network.

Of the four counselors, two had a graduation degree in basic science and 2-5 years of experience in health communication. One counsellor was a trained dietician and the remaining counsellor had extensive training in early childhood development. The protocols designed by the team were reviewed by a public health specialist. The counselors received 25 days of training on nutrition, early childhood development and child health from inhouse professionals, resource persons from Nutritional Rehabilitation and Research Centre, LTMG Hospital, Mumbai which is the nodal government centre for nutritional rehabilitation in Mumbai and NGOs such as Foundation for Mother and Child Health and Ummeed Child Development Centre.

Calls were made from mobile phones with dedicated lines. A web-based application was used to document the details of all attempts made to call each enrolled participant. A paper-based system was used to schedule and track calls. As per protocol, a call was scheduled on the same day and time of the week according to the preference of the participant. However, counsellors made ad hoc adjustments to accommodate the needs of the participants. As a rule, all calls to a participant were delivered by the same counsellor. In exceptional cases, calls were transferred to another counsellor for a temporary period. Whenever possible, participants were informed beforehand of this change.

## CONDUCT OF THE STUDY

An in-house field team of 8 fieldworkers was rigorously trained to conduct the fieldwork. These fieldworkers had 2-3 years of prior experience in monitoring child health and using weighing scales. They were given extensive training in data collection, informed consent procedures and the use of the mobile application used to record the data. Two field supervisors undertook the monitoring and evaluation activities of the trial. A house-to-house survey was conducted and all eligible infants were weighed. Analog weighing machines were used to record weight. The date of birth of the child was verified by reviewing available medical records. Physical growth monitoring charts were used for classification. Infants who were classified as moderately underweight were further screened for underlying comorbidities. The primary caregiver of those who met the inclusion criteria were enrolled in the study after seeking written informed consent. The consent was sought by the trained field investigators A baseline survey was conducted to obtain socio-demographic information, details of the child’s health history, dietary intake, feeding and caregiving, phone access and digital literacy.

As per protocol, an endline assessment was planned to be conducted six months after the conduct of the baseline interview. The endline assessment involved obtaining the current weight of the child and assessment of present knowledge and behaviour in relation to best practices in infant and young child feeding, health and hygiene. However, due to reasons such as relocation of the household, visits to the native place and unavailability on the day of the visit, there was variability in the recall period. The median time gap between the baseline and endline assessments was 9 months.

### Sample description and profile

As per the sample size calculation, we screened 1572 infants till we were able to identify 800 eligible participants. After excluding invalid phone numbers, incomplete forms and ineligible beneficiaries, we had enrolled 785 participants. Of these, 388 were assigned to the control arm and 397 to the intervention arm. An endline assessment could be conducted with 713 (91.0 percent) participants. The loss to follow up was similar across both arms, being 37 and 35 in the control and intervention arms respectively (chi square = 1.331; p = 0.249). (Figure 1)

The socio-demographic profile of the mothers who were surveyed at endline was similar across both the groups **(Table 1)** with no statistical difference in mother’s education, father’s occupation or birth order of the index child. The proportion of mothers having less than 10th standard education was 44 percent in both arms. The proportion of unskilled workers, including unemployed workers was 76 percent in the control group and 80 percent in the intervention group. Across both arms, in 33 percent of the cases, the index child was the first child.

**Table 1:** Background characteristics of the participants re-surveyed at endline.

| Characteristics | Control | Intervention | Chi square | P Value |
| --- | --- | --- | --- | --- |
| **Completed Educational level of mother** |  |  |  |  |
| Less than 10th grade | 151 (44.3%) | 155 (44.0%) | 0.007 | 0.939 |
| 10th grade or higher | 192 (55.7%) | 197 (56.0%) |  |  |
| Data Missing | 8 | 10 |  |  |
| **Occupation of the father/main earner** |  |  |  |  |
| Unemployed/Unskilled Worker | 268 (76.4%) | 289 (80.1%) | 1.432 | 0.239 |
| Skilled worker/service sector worker | 83(23.6%) | 72(19.9%) |  |  |
| Data Missing | 0 | 1 |  |  |
| **Birth order of index child** |  |  |  |  |
| First child | 116 (33.2%) | 118 (32.7%) | 0.24 | 0.936 |
| Second or later child | 233 (66.8%) | 243(67.3%) |  |  |
| Data Missing | 2 | 1 |  |  |

The age of the mothers receiving counselling and of the infants were also similar across the control and intervention arms with no statistical difference **(Table 2)**. The median age of the intervention group was 26 years as opposed to 25 years for the control group. The mean age was 27 years for both groups. The median age of the infants in the intervention group at baseline was 14 months as compared to 16 months for the control group. While all the enrolled infants were classified as moderately underweight, there was a slight difference in the mean weight at baseline between the intervention group infants (7.50 kgs) and the control group (7.59 kgs) which was consonant with the difference in their median age at baseline. This difference was not statistically significant.

**Table 2:** Age and weight characteristics of mother and index child.

| Characteristics of the mother and index child |  |  |  |  |
| --- | --- | --- | --- | --- |
|  | Control | Intervention | F | P value |
| Mean age of mother at baseline | 27 years | 27 years | 0.055 | 0.814 |
| Median age of mother at baseline | 25 years | 26 years |  |  |
| Mean age of the index child at baseline | 16 months | 15 months | 0.472 | 0.492 |
| Median age of the index child at baseline | 16 months | 14 months |  |  |
| Mean weight of the index child at baseline | 7.84 kgs | 8.27 kgs | 0.844 | 0.359 |
| Median weight of the index child at baseline | 8 kgs | 8 kgs |  |  |

Within the broad inclusion criteria of moderately underweight < -2SD and > -3SD, all infants were categorised by percentile categories as per the WHO growth standards (30). As the outcome of interest was change in the pattern of weight gain over a relative short period, we chose percentile category to analyse results. At baseline, the distribution of infants was very similar across the percentile categories was very similar. About two thirds of the infants were in the lowest one percentile. Almost all the infants occupied positions in the lowest 3 percentiles.

**Figure 3:**
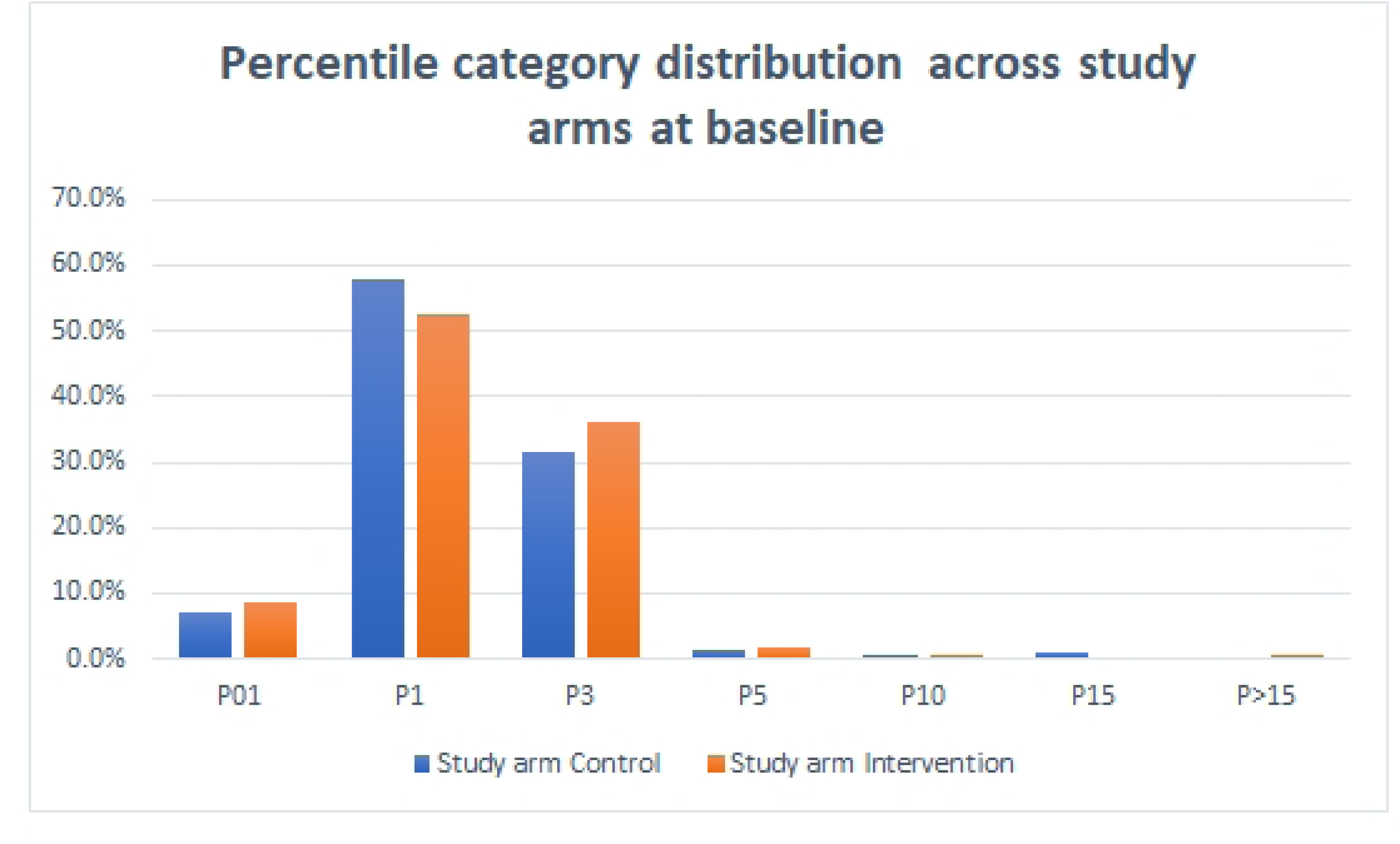
Distribution of infants by percentile category at baseline.

## RESULTS

### Exposure to the counselling calls

As noted earlier, calls were initiated to all participants assigned to the intervention arm. Among the 362 participants who also completed the endline evaluation, 274 (76 percent) had received at least one call **(Table 3)**. 51 percent of the participants had received 5 or more calls. Among the 274 participants who had received at least one call, the median number of calls was 9. The median duration of a counselling call was 6 minutes. For the beneficiaries who attended at least one call, the median duration of engagement (total duration of all successful calls) was 47.5 minutes. For those whom we categorised as engaged beneficiaries (attended at least five calls or more), the median duration of engagement was 55 minutes.

**Table 3:**
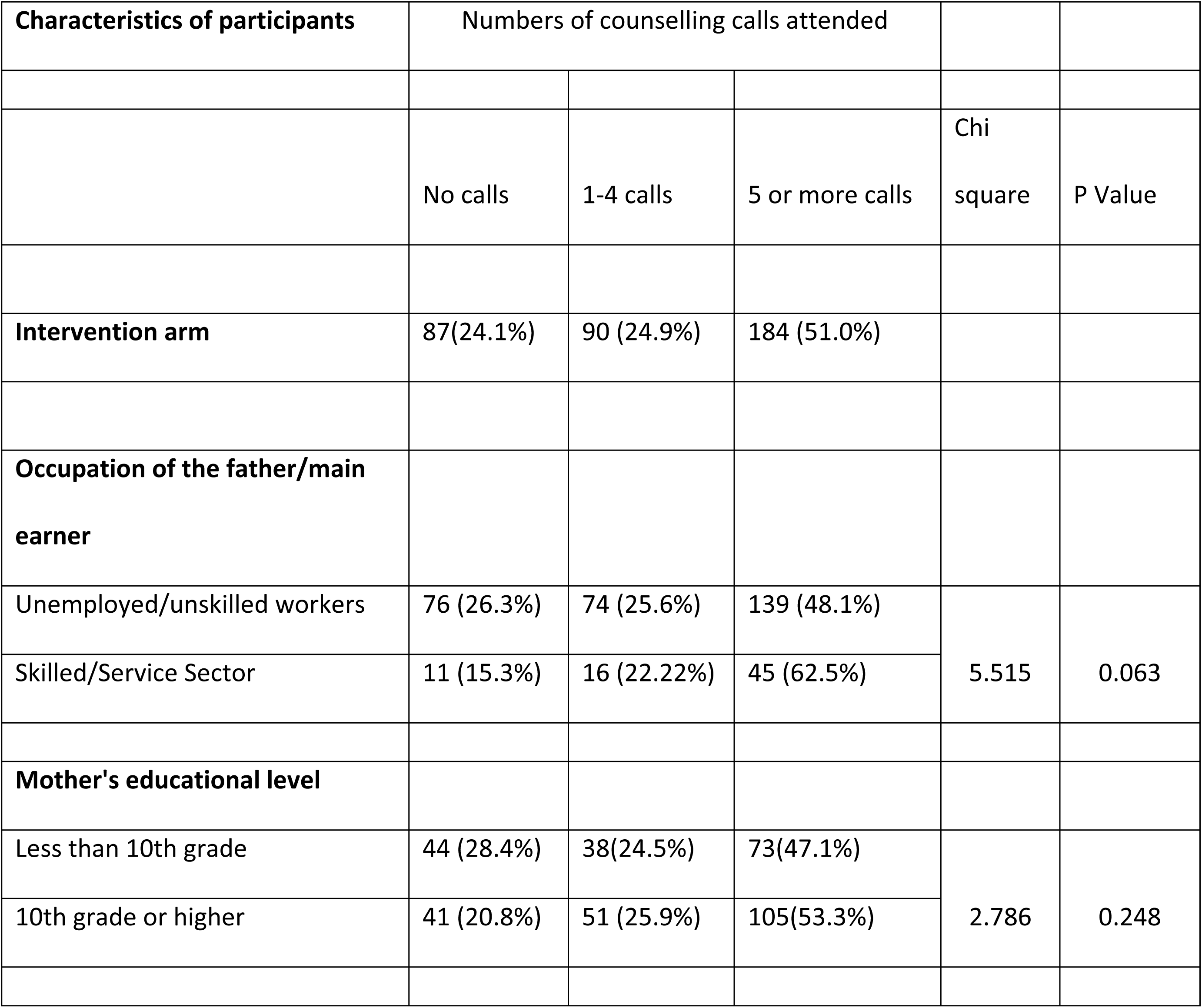
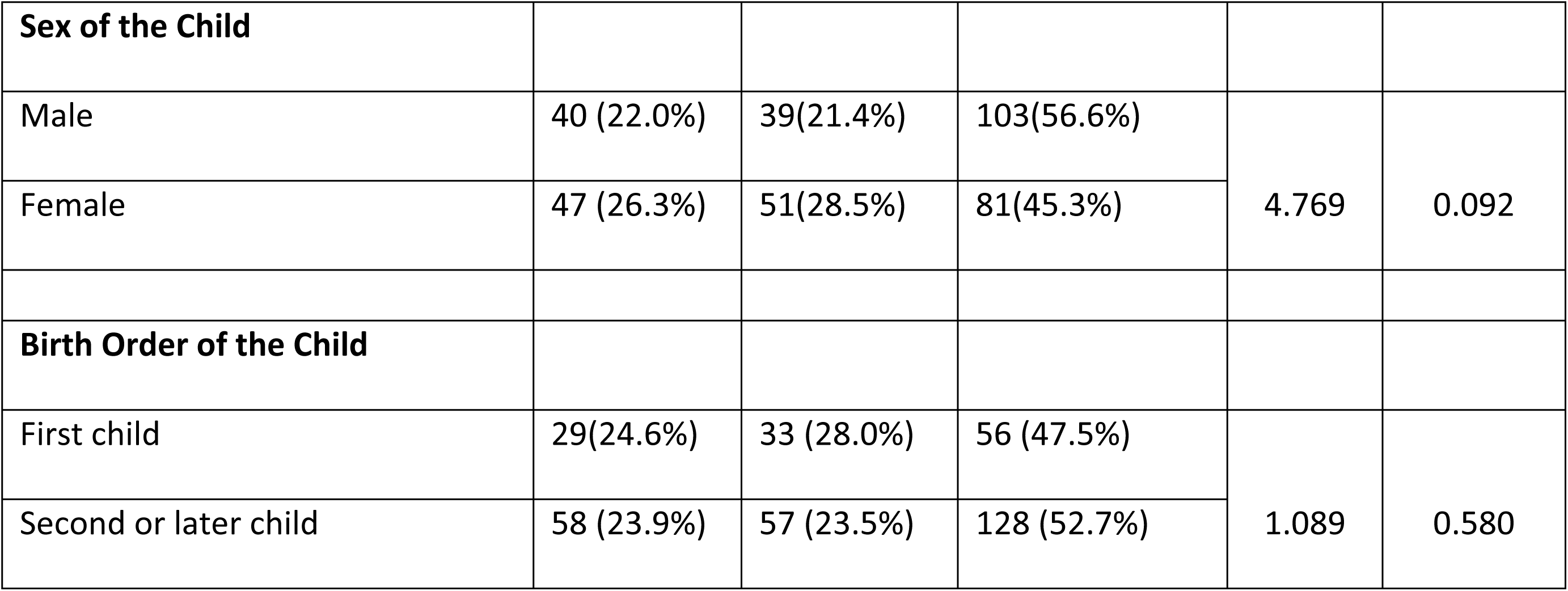
Number of calls attended by characteristics of the participants.

| Characteristics of participants | Numbers of counselling calls attended |  |  | Chi square | P Value |
| --- | --- | --- | --- | --- | --- |
|  | No calls | 1-4 calls | 5 or more calls |  |  |
| **Intervention arm** | 87(24.1%) | 90 (24.9%) | 184 (51.0%) |  |  |
| **Occupation of the father/main earner** |  |  |  |  |  |
| Unemployed/unskilled workers | 76 (26.3%) | 74 (25.6%) | 139 (48.1%) | 5.515 | 0.063 |
| Skilled/Service Sector | 11 (15.3%) | 16 (22.22%) | 45 (62.5%) |  |  |
| **Mother's educational level** |  |  |  |  |  |
| Less than 10th grade | 44 (28.4%) | 38(24.5%) | 73(47.1%) | 2.786 | 0.248 |
| 10th grade or higher | 41 (20.8%) | 51 (25.9%) | 105(53.3%) |  |  |

| **Sex of the Child** |  |  |  |  |  |
| --- | --- | --- | --- | --- | --- |
| Male | 40 (22.0%) | 39(21.4%) | 103(56.6%) | 4.769 | 0.092 |
| Female | 47 (26.3%) | 51(28.5%) | 81(45.3%) |  |  |
| **Birth Order of the Child** |  |  |  |  |  |
| First child | 29(24.6%) | 33 (28.0%) | 56 (47.5%) | 1.089 | 0.580 |
| Second or later child | 58 (23.9%) | 57 (23.5%) | 128 (52.7%) |  |  |

There were interesting trends in the patterns of engagement. The mother’s educational level did not have a very significant influence on engagement. While women who had completed school were more likely to attend 5 or more calls (53.3 percent) than those who had not completed school (47.1 percent), the difference was not statistically significant. Similarly, women who were first time mothers were a little less likely to attend five or more calls (47.5 percent) than those with more than one child (52.7 percent). However, this difference was also not statistically significant. However, the sex of the child had a more visible effect on engagement, with the mothers of boys were likely to attend 5 or more calls (56.6 percent) as compared to the mothers of girls (45.3 percent). The mothers whose husbands/heads of households were unskilled workers had lower engagement with more than 1/4^th^ (26.3%) not engaging at all.

### Primary outcome: Nutritional status at endline

The study had an intention to treat design with a focus on improving the nutritional status of the child through improved dietary, health and hygiene practices. As the study had a short recall period, we focused on changes in the trajectory of growth which were measured using three different metrics. These metrics indicated both absolute as well as relative change in nutritional health as defined by the WHO Growth Standards. Weight for age z scores (WAZ) are accepted universally as measure of nutritional health and were also used in this study to define the inclusion criteria. Thus, we analysed the nutritional status of the infants at endline and classified them into the three categories of normal weight (> - 2SD), moderately underweight (< - 2SD and > - 3SD) and severely underweight (< - 3SD) **(Table 4)**. A similar proportion of infants had recovered in both arms (22.2% in control and 22.7% in intervention arm). However, the proportion of infants who had declined to severe underweight was significantly lower in the intervention arm, being 7.5% as against 11.5% in the control arm. The difference was not statistically significant.

**Table 4:** Nutritional status of infants at endline by study arm.

| Normal Weight (WAZ $> -2SD$) | | Moderately Underweight (WAZ $< -2SD$ & $> -3SD$) | | Severely Underweight (WAZ $< -3SD$) | | Total | |
| --- | --- | --- | --- | --- | --- | --- | --- |
| Control | Intervention | Control | Intervention | Control | Intervention | Control | Intervention |
| 78 | 82 | 232 | 253 | 41 | 27 | 351 | 362 |
| 22.2% | 22.7% | 66.1% | 69.9% | 11.7% | 7.5% | 100.0% | 100.0% |

We also compared the results using percentile category defined by the World Health Organisation’s child growth standards, which are specific for age and sex. In both arms, the distribution of infants was similar at baseline. A Mann-Whitney U test results indicated no statistically significant difference in the distribution of percentile categories between the control and intervention groups, (U=64481.5,p=.541).

However, at the endline, we find that fewer infants in the intervention arm had remained in the lowest percentile category as compared to the control arm. While 34 percent of the infants in the control arm were in the lowest 1 percentile, this figure was 26 percent for the intervention arm **(Figure 4).** However, though there were observable differences in distribution, they were not statistically significant ((U=68209.0; p=.071).

**Figure 4:**
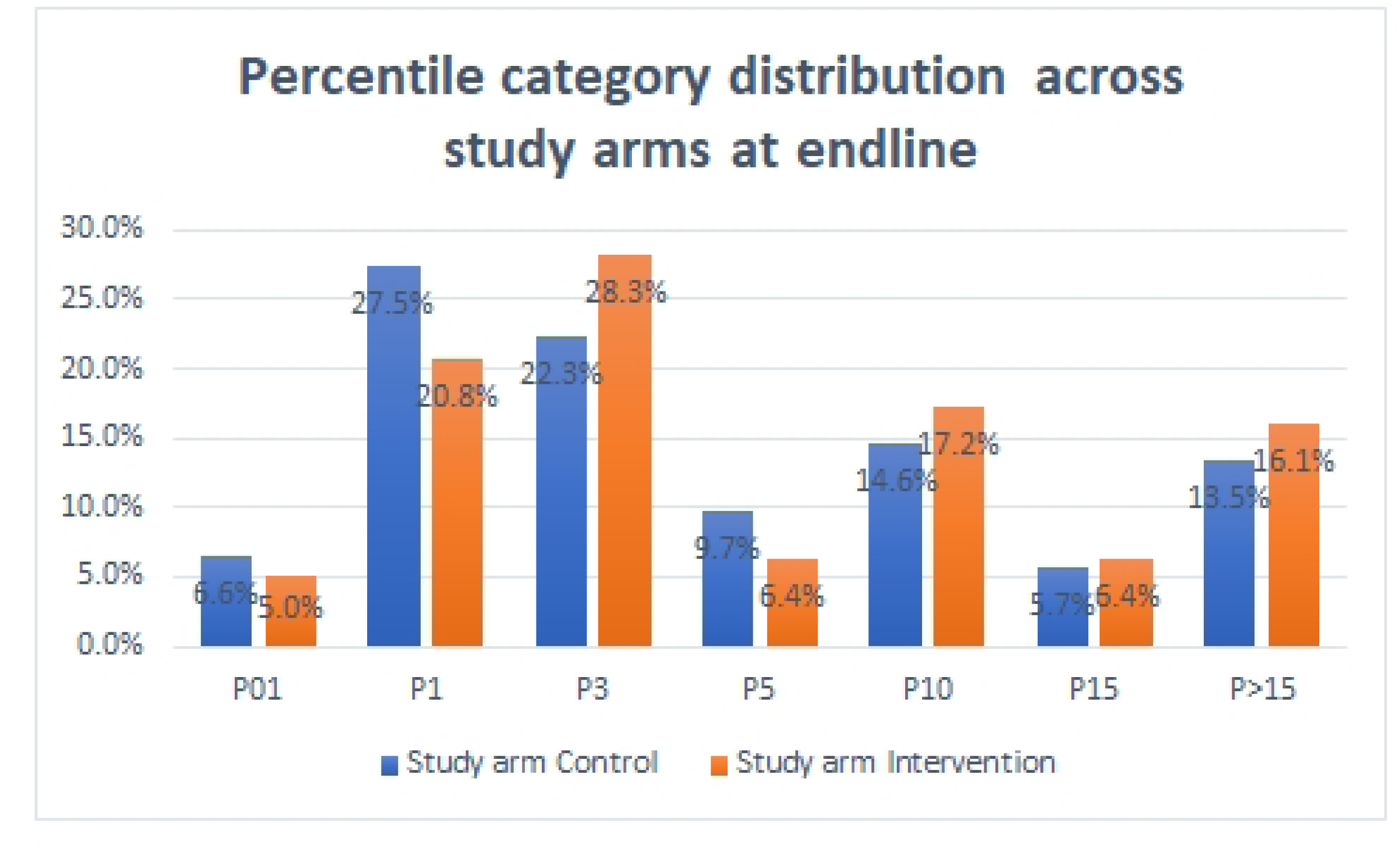
Percentile category Distribution of infants across study arms at endline.

The overall results did not indicate any statistical difference between the study arms at endline in the change in nutritional category or percentile category, although there was a visible trend of greater improvement in the intervention arm.

We conducted a second level of analysis at the individual level, to assess improvement in nutritional health relative to their own status at baseline. A difference in difference analysis was conducted by tracking changes in the percentile category at baseline and endline for each child. Movement to a higher percentile category was classified as ‘improvement’, whereas movement to a lower category was classified as deterioration. In cases, where the percentile category remained the same, it was assumed that there was no change in the rate of weight gain. While there was an improvement in 59.5 percent of the infants in the control group, this figure was 67.1 percent in the intervention group. Simultaneously, the proportion of infants who moved to a lower percentile category was 12.8 percent in the control arm as opposed to 10.5 percent in the intervention arm. **(Table 5).** However, this difference was not statistically significant (Chi square = 4.417; p value = 0.110)

**Table 5:** Difference in difference analysis of percentile category position at baseline and endline of infants by intervention arm.

| Percentile Change | Control | Intervention | Grand Total |
| --- | --- | --- | --- |
| Improvement | 209 (59.5) | 243 (67.1) | 452 |
| No Change | 97 (27.6) | 81(22.4) | 178 |
| Deterioration | 45 (12.8) | 38 (10.5) | 83 |
| Grand Total | 351 | 362 | 713 |

It is well understood that childhood malnutrition disproportionately affects infants living in poverty (33). Household diets are influenced by gender norms and culture (34). Particularly in India, there is gender disparity in the access to food as well as health care resources. (35). Hence, we disaggregated the results for social groups, which face higher social risk even within the largely disadvantaged population included in the study to assess if there were differing trends across sex, social class and education. The results indicated that there were starkly differing trends. **(Table 6)**. The female infants in the intervention group fared much better than their counterparts in the control group. Only about half as many deteriorated (7.8% v/s 14.3%) and significantly more improved (76.0% v/s 64.0%). Strikingly, for male infants, the difference between the study arms was negligible. Likewise, for poorer infants, who lived in households of unskilled workers, there was 10 percentage point increase in improvement and 4 percentage point lowering in deterioration. The differences were statistically significant. In comparison, among infants from the better off households, there was no difference in the outcomes at endline.

**Table 6:** Relative change in percentile category position at endline for infants facing higher health risk or higher social risk across intervention and control arms.

|  |  | Improvement | No change | Deterioration | Chi Square | Significance |
| --- | --- | --- | --- | --- | --- | --- |
| Sex of the infant |  |  |  |  |  |  |
| Female | Control | 64.0% | 21.7% | 14.3% | 6.590 | p = 0.037* |
|  | Intervention | 76.0% | 16.2% | 7.8% |  |  |
| Male | Control | 54.6% | 33.9% | 11.5% | 1.228 | p = 0.541 |
|  | Intervention | 58.2% | 28.6% | 13.2% |  |  |
| Occupation of main earner |  |  |  |  |  |  |
| Unskilled Workers | Control | 57.5% | 29.1% | 13.4% | 6.033 | P=0.049* |
|  | Intervention | 67.5% | 22.8% | 9.7% |  |  |
| Service/Skilled Workers | Control | 66.3% | 22.9% | 10.8% | 0.372 | P = 0.830 |
|  | Intervention | 65.3% | 20.8% | 13.9% |  |  |

Among infants who had low birth weight, almost half (48 percent) had no change or deteriorated in terms of the percentile category of weight in the control group **(Table 7)**. In contrast, only 28 percent of such infants in the intervention group had failed to improve. The difference in outcomes was statistically significant. Infants who were placed in the lowest percentile category at baseline (P0.01 and P1) showed a higher probability of improvement in both arms. However, the proportion of infants with no change or deterioration in percentile category position was significantly lower for infants in the intervention arm being 26 percent as opposed to 37 percent in the control arm. This difference was also statistically significant.

|  |  | Improvement | No<br>change | Deterioration | Chi Square | Significance |
| --- | --- | --- | --- | --- | --- | --- |
| Birth Weight of Infant |  |  |  |  |  |  |
| Low Birth Weight | Control | 51.9% | 36.7% | 11.4% | 8.402 | 0.015* |
|  | Intervention | 72.5% | 21.1% | 6.4% |  |  |
| Normal Birth Weight | Control | 61.4% | 25.4% | 13.3% | 0.419 | 0.811 |
|  | Intervention | 64.1% | 23.3% | 12.7% |  |  |
| Percentile Category at<br>baseline |  |  |  |  |  |  |
| Lowest 1 percentile (P01, P1) | Control | 63.0% | 30.4% | 6.6% | 6.786 | 0.034* |
|  | Intervention | 74.3% | 21.6% | 4.1% |  |  |
| Higher than lowest<br>percentile (P2 and above) | Control | 53.2% | 24.2% | 22.6% | 0.459 | 0.795 |
|  | Intervention | 55.7% | 23.6% | 20.7% |  |  |

## DISCUSSION

The study attempted to evaluate the outcomes of offering advice and support to caregivers of moderately underweight infants via telephonic calls. This live telephonic counselling intervention was aimed at bridging the gap in the existing services which does not focus on moderate malnutrition and does not emphasise counselling. While there are several sources of nutritional advice, an attempt was made in this intervention to proactively approach mothers and give them an opportunity for receiving individualised inputs. The medium selected for the intervention deployed the most basic technology (live calls) which is widely available and accessible to all women. In this paper, we have reported the results pertaining to the primary outcome, i.e. accelerated weight gain and reduction in decline to more severe levels of malnutrition.

The results indicate that the intervention was quite successful in achieving its primary outcome of accelerated weight gain. For the control and intervention arms as a whole, the probability of infants moving to a higher percentile category was higher for the latter. The more accentuated difference for infants who were at higher levels of social risk on account of gender and economic disadvantage indicated the potential of the intervention to foster greater equity. Significantly, for infants who were already at a higher risk of severe malnutrition being born low-birth weight or having weight in the lowest percentile category, the relative gains were more significant. The rather negligible effect for the other groups point to the need for better targeting.

The results of this study reiterate the fact that there is an important role for counselling in nutritional health. Other studies evaluating on-ground counselling programmes have demonstrated improvement in weight for age (33). A systematic review of home/community-based nutrition education studies indicates the need for specialised interventions for malnourished children in contrast to the generic education provided to all mothers (34). This is arguably one of the first systematic studies to demonstrate the results of tele-counselling for addressing childhood malnutrition by delivering individualised advice.

## CONCLUSION

Notably, the gains were highest for those infants (female infants, those have a father in unskilled occupations) despite the fact that, as a group, their caregivers had lower engagement. This indicates that the barriers for these groups to access the programme were greater, as against a likelihood of greater benefit as well. Arguably, measures to prevent their dropout from the programme would result in even greater gains. This points to the need to improve access to technology and addressing social/cultural barriers as a precursor for implementing an mhealth programme. Subsequent to this study, there were several changes which were instituted in the programme to consciously address gender bias, both among the participants of the programmes as well as the care-workers in the team. Better targeting was also institutionalised to ensure that more efforts were made to engage with and retain more marginalised participants. The results of this study indicate a potential for including telephonic counselling as an integral part of nutrition programmes for children across diverse settings.

## Data Availability

The data is owned wholly by ARMMAN. It is not available in the public domain. Deidentified and unlinked data may be made available to third parties on request in compliance with the data sharing policy of ARMMAN and as outlined in the bilateral data sharing agreement with the requesting party.

## Limitations of the study

This study is limited to one Indian metropolitan city and studied outcomes over a short period. Hence, it is possible that the peculiarities of the health system and characteristics of the population would imply that these results have limited generalisability. However, there were definite indications that the intervention has positive results and is particularly effective for populations and infants with more vulnerability in terms of socio-economic status, age, sex and severity of malnutrition.

## Ethical Considerations

The study was reviewed and approved by the ARMMAN Ethics Review Board. The study was registered as an interventional trial with the Clinical Trials Registry of India (CTRI/2023/01/048939) with the title “Improving adherence to best practices in feeding, health and hygiene amongst moderately underweight children through telephonic counselling “

## Acknowledgements

We would like to acknowledge the entire field team and counsellors of Swasth Kadam, who work tirelessly for conducting this study. We would like to thank Komal Gholekar for her support for data management. We would like to particularly acknowledge Dr. Anuja Jayaraman’s support for reviewing successive drafts of this paper.

## Contributorship statement

Authors, Neha Madhiwalla, Hemlata Jiwnani and Bharati Jadhav were involved in the designing and implementation of the study. Hardik Bhatt was involved in the analysis of data. Aparna Hegde was involved in the designing of the study. All the authors contributed to writing of the manuscript.

